# Genetic variants associated with longevity in long-living Indians

**DOI:** 10.1101/2024.04.19.24305679

**Authors:** Sandhya Kiran Pemmasani, R G Shakthiraju, V Suraj, Raunaq Bhattacharya, Chetan Patel, Anil Kumar Gupta, Anuradha Acharya

## Abstract

Genetic factors play a significant role in determining an individual’s longevity. The present study was aimed at identifying the genetic variants associated with longevity in Indian population. Long living individuals (LLIs), aged 85+, were compared with younger controls, aged 18-49 years, using data from GenomegaDB, a genetic database of Indians living in India. An in-house developed custom chip, having variants associated with various cancers, cardiovascular, neurological, gastro-intestinal, metabolic and auto-immune disorders, was used to generate genotype data. Logistic regression analysis with top five genetic principal components as covariates resulted in 11 variants to be significantly associated with longevity at a p-value threshold of 5×10^−4^. Alleles associated with slower heart rate (rs365990, MYH6), lower risk of osteoporosis and short body height (rs2982570, ESR1), decreased risk of schizophrenia (rs1339227, RIMS1-KCNQ5) and decreased risk of anxiety and neuroticism (rs391957, HSPA5) were found to have higher frequency in LLIs. Alleles associated with increased risk of atrial fibrillation (rs3903239, GORAB-PRRX1) and biliary disorders (rs2002042, ABCC2) were found have lower frequency. The G allele of rs2802292 from FOXO3A gene, associated with longevity in Japanese, German and French centenarians, was also found to be significant in this population (P=0.027). Pathway enrichment analysis revealed that the genes involved in oxidative stress, apoptosis, DNA damage repair, glucose metabolism and energy metabolism were significantly involved in increasing the longevity. Results of our study demonstrate the genetic basis of healthy aging and longevity in the population.

## INTRODUCTION

Longevity and healthy ageing are complex phenomena influenced by various factors, like socioeconomic status, nutrition, lifestyle, physical activity, gender and genetics. Among them, genetics plays a major role, and its influence is estimated to be around 25-40 percent [1]. Studies indicate that offspring of long-lived individuals tend to lead a healthy and longer life compared to general population [2]. This could be due to inheritance of genetic variants associated with healthy lipid profiles, enhanced insulin sensitivity, slower cognitive decline and lower incidence of age-related diseases, like Alzheimer’s and cardiovascular diseases [3-7]. There are also a certain category of variants that are identified to be prevalent in long-living individuals through multiple genome-wide association studies (GWAS), candidate studies and meta-analyses related to longevity. They are the variants from Forkhead Box O3A (FOXO3A) and Apolipoprotein E (APOE) genes that are part of pathways associated with cell apoptosis, metabolism and oxidative stress [8-10]. Variants that decrease the length of telomeres also influence an individual’s lifespan [11-12].

Genetic associations observed in one population might not reflect in other populations due to variable frequencies of effect allele across populations. In developed countries or high-income countries, reaching 100 years of age is easier compared to low-income countries, and hence selection pressure is less. This leads to relatively lesser frequencies of effect alleles [13]. Also, age thresholds to define “cases” or “long-living individuals” change as per the population under study. Studies done on isolated populations (Blue Zones) show that the environmental factors work in synergy with genetics in contributing to exceptional longevity in those populations [14]. So, for any study done on genetics of longevity, it is necessary to understand longevity of the individuals in that population, define the age thresholds and take cases and controls from the same environment.

According to the latest estimates of Longitudinal Aging Study in India (LASI - 2019), India has world’s second-largest number of older people, aged 60 and above [15]. It is further estimated that the number will double by 2050. With improved access to vaccines, antibiotics and clean water, and increased awareness on healthy diets and physical activity, life expectancy at birth has improved from 62.1 years in 2000 to 70.8 years in 2019 [16]. Despite this, only 0.4% of the population reaches the age of 85 years. Understanding the genetics of these long living individuals (LLIs), of age 85+, in India is still a challenge due to lack of genetic research on the population. It is necessary to conduct genome-wide studies by selecting cases and controls from the individuals living in India. Identifying genetic variants associated with longevity, and in turn inferring factors that influence longevity helps in recommending strategies for healthy aging at population-wide and an individual level.

In the present study, we have identified variants that have significantly higher frequency in long-living Indians, compared to younger controls, using GenomegaDB of Mapmygenome [17]. Through literature mining on the identified variants we have inferred the factors that help in healthy aging. To our knowledge this is the first genetic study to investigate variants associated with longevity in Indian population.

## MATERIALS AND METHODS

### Study Data

Participants in the current study were taken from GenomegaDB of Mapmygenome, which is a genotype-phenotype database of Indians living in India. Cases were defined as participants with self-reported age of greater than 85 years at the time of sample collection. Controls were the participants with 18-49 years of age at the time of sample collection. A total of 133 cases and 1155 controls were considered for the present study, to investigate the variants associated with longevity in Indian population. Genotyping was done using a custom chip developed in-house at Mapmygenome. It has 8,768 variants associated with various phenotypes, like cancers, cardiovascular, neurological, gastrointestinal, metabolic and autoimmune disorders. Written informed consent was taken from each participant. Research was carried out in compliance with the Helsinki declaration and the procedures had been approved by internal bio-safety committee at Mapmygenome.

### Data pre-processing

Variants with genotype call rate <90%, minor allele frequency (MAF) <5% and are not in Hardy-Weinberg Equilibrium (HWE) in controls (P_HWE_ <= 0.0001) were removed from further analysis. Also, the participants with genotype call rate <95% were removed from the analysis. In the case of relatives, only one participant, preferably the elder one was retained in the analysis. Participants who were genetic outliers, defined as being more than three standard deviations away from the mean of top five genetic principal components, were excluded from the analysis. R [18] and PLINK [19] software were used for all the data pre-processing.

### Association analysis

The association of each variant with longevity was assessed using additive logistic regression with Firth’s bias reduction available in PLINK software. Top five genetic principal components were added as covariates in the model, to correct for population stratification. Variants with p-value <= 5×10^−4^ were considered to be significantly associated with longevity. Manhattan plots were generated using *ggplot* package of R v4.2. Gene information was obtained through NCBI’s E-utilities [20].

### Pathway enrichment analysis

Genes corresponding to the significant SNPs were mapped to KEGG (Kyoto Encyclopedia of Genes and Genomes) pathway maps to understand the possible functional processes associated with longevity. DAVID Functional Annotation Tool v2024q1 [21-22] and ClueGO v2.5.10 [23] were used to perform pathway enrichment analysis and to draw interacting networks.

## RESULTS

Table 1 gives information on characteristics of participants included in the analysis. Mean age of cases was found to be 93.4 years, and mean age of controls was found to be 36.4 years. There was equal representation of participants from both the genders. Out of 8,768 variants, 6,300 autosomal SNPs passed quality control filters, and were finally considered in the analysis.

**Table 1.** Characteristics of study participants.

|  | Cases | Controls |
| --- | --- | --- |
| Number of participants | 133 | 1155 |
| Age, mean (SD) | 93.4 (6.0) | 36.4 (7.7) |
| Male, n (%) | 65 (49%) | 647 (56%) |

Association analysis using additive logistic regression along with top five genetic principal components resulted in 11 variants to be significantly associated with longevity, at a p-value threshold of 5×10^−4^. Table 2 gives information on minor allele, major allele, minor allele frequency (MAF), p-value and odds ratio obtained for these variants. Figure 1 shows Manhattan plot of associations. 492 variants that have suggestive association at a nominal p-value of 0.05 are given in Supplementary Table 1.

**Table 2.** Variants significantly associated with longevity in Indian population.

| ID | Chr | Position | Minor allele | Major allele | MAF in Cases | MAF in Controls | P | OR | 95% CI | Gene/Nearest Gene |
| --- | --- | --- | --- | --- | --- | --- | --- | --- | --- | --- |
| rs12083278 | 1 | 31151182 | G | C | 0.149 | 0.084 | 3.125E-04 | 2.119 | 1.409 - 3.188 |  |
| rs1877455 | 1 | 114556471 | T | C | 0.297 | 0.433 | 1.489E-04 | 0.588 | 0.447 - 0.774 | TRIM33-BCAS2 |
| rs3903239 | 1 | 170600176 | G | A | 0.128 | 0.232 | 2.500E-04 | 0.498 | 0.343 - 0.723 | GORAB-PRRX1 |
| rs9363918 | 6 | 68432116 | T | G | 0.624 | 0.484 | 4.700E-05 | 1.716 | 1.323 - 2.226 | LINC02549-ADGRB3 |
| rs1339227 | 6 | 72445999 | T | C | 0.493 | 0.375 | 3.602E-04 | 1.604 | 1.237 - 2.079 | RIMS1-KCNQ5 |
| rs2982570 | 6 | 151692613 | T | C | 0.290 | 0.195 | 3.882E-04 | 1.692 | 1.265 - 2.262 | ESR1 |
| rs10970985 | 9 | 32453121 | C | G | 0.267 | 0.398 | 5.984E-05 | 0.56 | 0.422 - 0.743 | ACO1 |
| rs391957 | 9 | 125241745 | T | C | 0.417 | 0.311 | 4.831E-04 | 1.586 | 1.224 - 2.055 | HSPA5 |
| rs2002042 | 10 | 99828174 | T | C | 0.105 | 0.197 | 4.309E-04 | 0.487 | 0.326 - 0.727 | ABCC2 |
| rs2762051 | 13 | 50261579 | T | C | 0.150 | 0.071 | 3.220E-05 | 2.222 | 1.525 - 3.237 | DLEU1 |
| rs365990 | 14 | 23392602 | G | A | 0.365 | 0.263 | 3.965E-04 | 1.625 | 1.242 - 2.126 | MYH6 |

ID: rsID of variant; Chr: Chromosome number; Position: GRCh38 position; MAF in Cases: Frequency of minor allele in Cases; MAF in Controls: Frequency of minor allele in Controls; P: P-value calculated from logistic regression after adjusting for top five principal components; OR: Odds Ratio for minor allele; 95% CI: 95% Confidence Interval of Odds Ratio;

**Figure 1.**
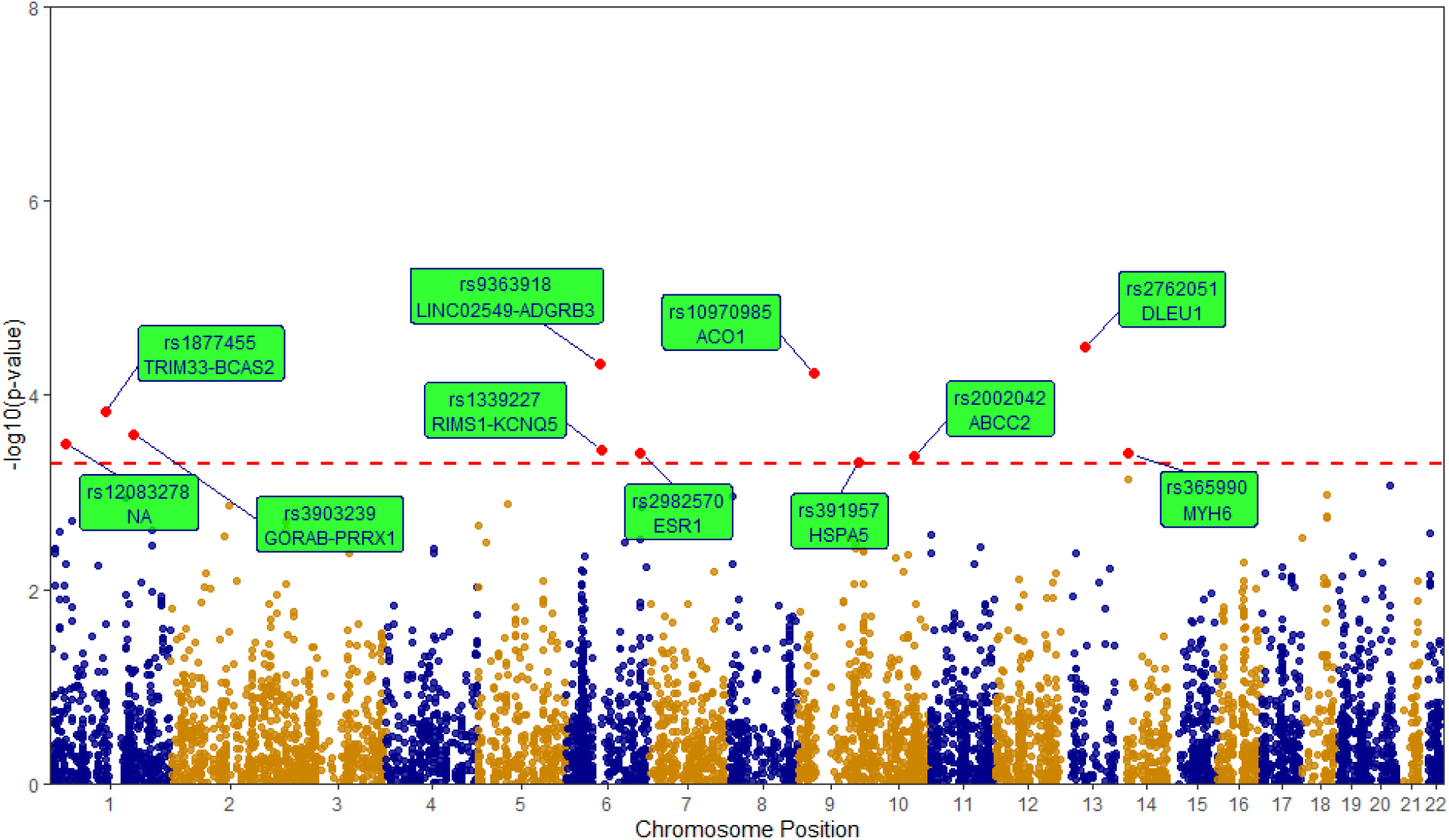
Manhattan plot of association analysis results.

Statistical significance of the association, as measured by the negative logarithm of the p-value, is shown on y-axis. Larger values correspond to smaller p-values. X-axis represents the chromosomal locations. Red horizontal dashed line indicates the p-value threshold of 5×10-4. Significant variants are highlighted in red, with their information in rectangular green boxes.

Literature mining on significantly associated SNPs revealed that they are implicated in various diseases, indirectly affecting the lifespan. As can be seen in Table 1, G allele of rs3903239, known to be associated with atrial fibrillation [24-25], decreases the chances of surviving to older age by 0.498 times (95% CI: 0.343 - 0.723). Its frequency is very low in LLIs compared to younger controls. Significant association was also observed for rs1339227, an intergenic SNP between RIMS1 and KCNQ5 genes. Several studies indicated that the C allele of this SNP is associated with schizophrenia [26-27]. Carriers of the other allele, T allele, at this locus have 1.604 times higher chances of surviving up to the age of 85 (95% CI: 1.237 - 2.079). The rs2982570, from ESR1 gene that is involved in growth and metabolism, is known to be associated with the risk of osteoporosis [28]. The non-effect allele, T allele, at this locus has an odds ratio of 1.692 (95%CI: 1.265 - 2.261) for longevity compared to the risk allele. The rs10970985, from ACO1 gene that is involved in controlling the iron levels in the cells, is found to have lower frequency of C allele in LLIs. The C allele at this locus decreases the survival to older age by 0.56 times (95% CI: 0.422 - 0.743). Another variant, rs391957 from HSPA5 gene, which is associated with decreased anxiety and neuroticism [29-30], was found to have high frequency in LLIs, with an odds ratio of 1.586 (95%CI: 1.224 - 2.055) for longer survival. Rs2002042 from ABCC2 gene, which is known to be associated with non-alcoholic fatty liver disease [31], has low frequency of risk allele in LLIs compared to controls. Presence of risk allele (T allele) decreases the chances of survival till 85+ by 0.486 times (95%CI: 0.326 - 0.727). Significant association with longevity was also observed for rs365990 from MYH6 gene that is involved in cardiac muscle contraction. G allele at this locus is known to be associated with prolonged PR interval and slower heart rate [32-33]. Carriers of G allele have 1.625 times higher chances of surviving to older age (95%CI: 1.242 - 2.126) than the carriers of A allele. The additional four SNPs in Table 1, rs12083278 [C] associated with severity of COVID-19 [34], rs1877455 [T] associated with autism along with other SNPs as haplotype [35], rs9363918 [T] associated with pancreatic cancer [36], and rs2762051 [T] associated with celiac disease [37], have limited literature support for their associations, and further research studies are required to understand their role in increasing the lifespan.

Previous studies showed multiple SNPs from FOXO3A gene to be associated with longer lifespan. In particular, the G-allele of rs2802292 showed significant presence in Japanese, German and French centenarians [38-40]. In our study, this SNP reached nominal significance with a p-value of 0.027 and an OR of 1.34. Other SNPs from FOXO3A (rs9400239, rs2153960, rs4946935 and rs4946936) did not show statistical significance, but the direction of association is consistent with other studies. Figure 2 shows the results of association analysis on SNPs from FOXO3A gene.

**Figure 2.**
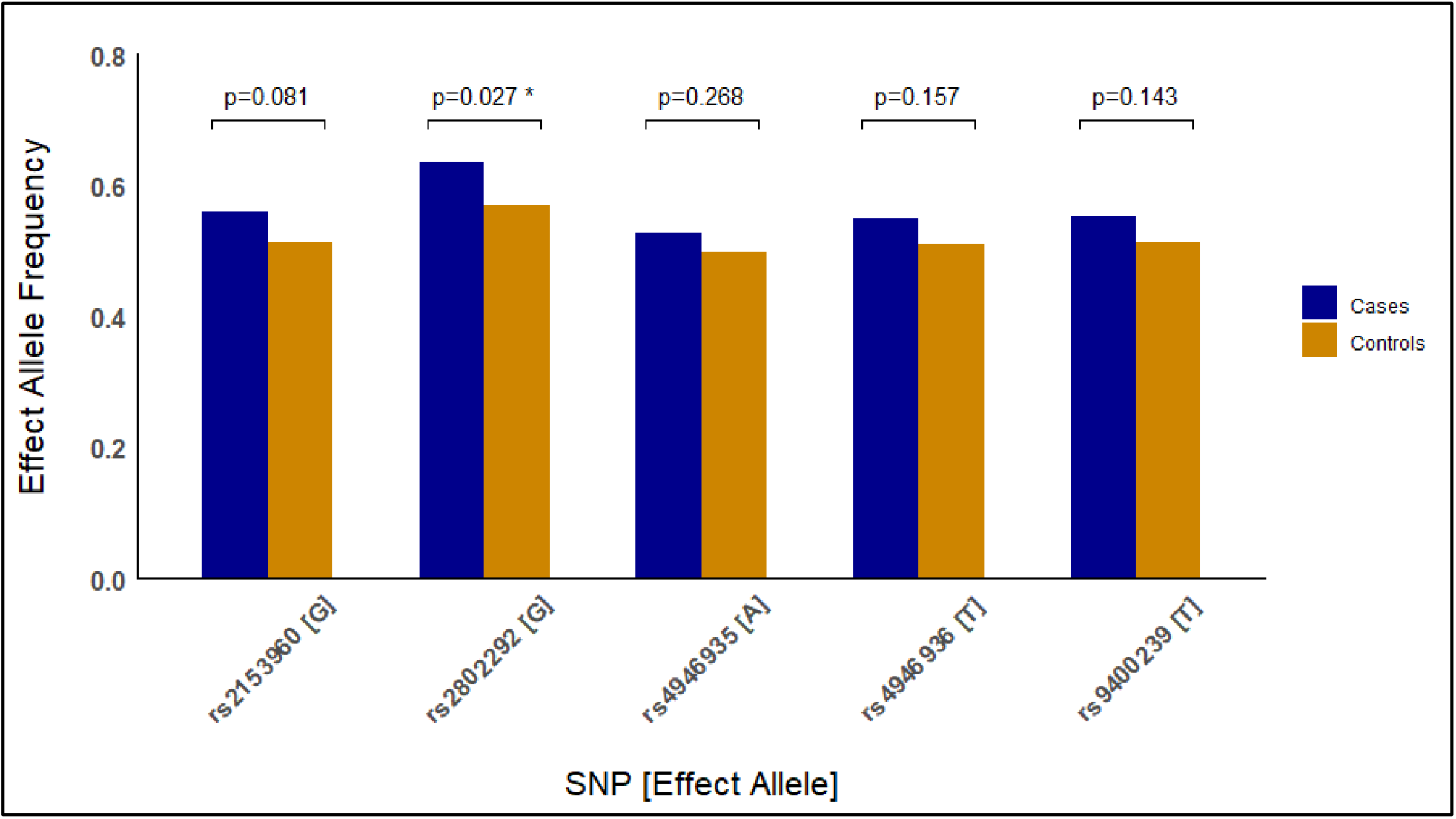
Association analysis on SNPs from FOXO3A gene.

Pathway enrichment analysis was performed on 492 variants that have suggestive association with longevity (p-value < 0.05). 84 variants located in intergenic regions did not map to any genes, and were excluded from the analysis. Significantly enriched KEGG pathways identified through DAVID Functional Analysis tool, with p-value threshold of 0.05, are given in Supplementary Table 2. Figure 3 shows the most significant pathways identified through ClueGO (P-value < 5×10^−4^). Significant pathways identified by both the tools were longevity regulating pathway, FoxO signaling pathway, adipocytokine signaling pathway and non-alcoholic fatty liver disease pathway. Some of the other pathways identified by the tools included insulin resistance, cholesterol metabolism, inflammatory bowel disease and PI3K-Akt signaling pathway.

**Figure 3.**
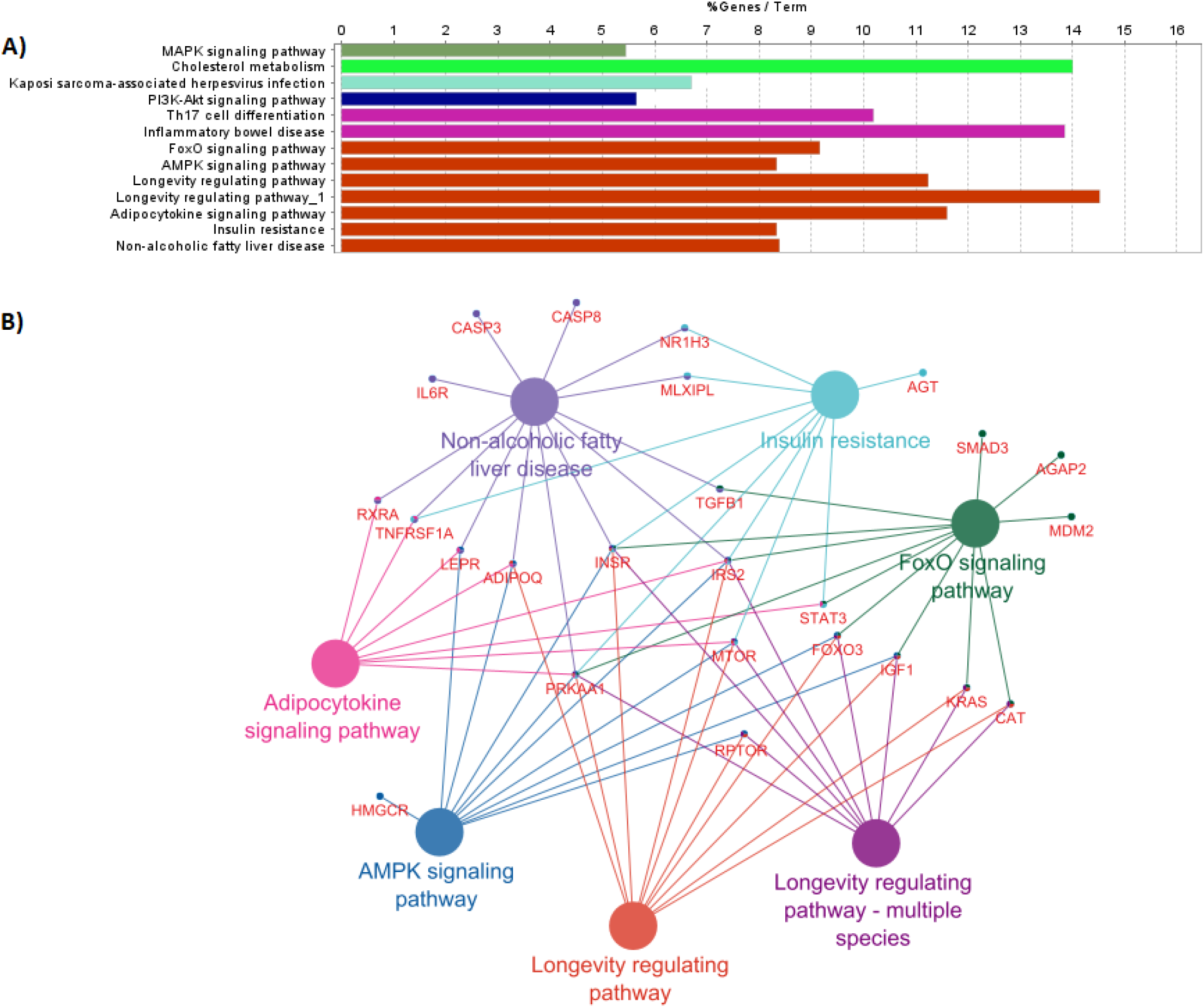
Pathway analysis through ClueGO. A) Barograph showing the % of genes mapped to each pathway term. Colors indicate the grouping of pathways done based on Kappa score B) Network of pathways grouped along with Longevity regulating pathway.

## DISCUSSION

In the present study, we analyzed genotype data of long-living Indians, aged 85+, to identify genetic polymorphisms associated with longevity in the population. By mapping the identified variants to genes, pathways and their functions, and by doing thorough literature search we inferred the factors that influence longevity and healthy aging. The alleles that increase the risk of atrial fibrillation, biliary disorders, schizophrenia, anxiety and neuroticism were in low frequency in long-living individuals compared to younger controls. Alleles associated with slower heart rate, lower risk of bone fractures, short body height were in high frequency.

Atrial fibrillation is one of the risk factors that reduce the life expectancy. It contributes to complications such as stroke and heart failure. Several genetic research studies indicated the association of rs3903239, an SNP in the upstream of PRRX1 gene, with atrial fibrillation. PRRX1 (Paired related homeobox 1) regulates muscle creatine kinase and controls the growth and development of mesodermal muscles such as the heart. SNPs in the upstream of PRRX1 may reduce the expression of the gene, and shorten the atrial action potential duration [41]. Ellinor et al [24] conducted a GWAS meta-analysis in individuals of European ancestry, and found G allele of rs3903239 to be significantly associated with atrial fibrillation. The association was also observed in Japanese and Korean population [25]. In the present study we observed a lower frequency of G allele in LLIs, indicating the lower risk of cardiovascular disorders, and increased longevity.

The T allele of rs1339227, which decreases the risk of schizophrenia, is present in higher frequency in LLIs. Schizophrenia Working Group of the Psychiatric Genomics Consortium identified significant association of rs1339227 with schizophrenia [26]. The T allele was found to decrease the risk of disease with an odds ratio of 0.942. Yao et al [27] found a decreased effect of T allele in the association of schizophrenia. Higher frequency of T allele in LLIs suggests protective association of this SNP in increasing the lifespan.

Estrogen receptor 1 gene (ESR1) encodes an estrogen receptor (ER-alpha) that plays a major role in bone metabolism and maintenance of the skeletal system. Several genome-wide association studies have found variations in this gene to be associated with osteoporosis and risk of fractures via low bone mineral density (BMD). Estrogen-replacement therapy in post-menopausal women is known to prevent bone loss and osteoporosis. Trajanoska et al [42] found the C allele of rs2982570 from ESR1 to be associated with increased risk of fractures and decreased femoral neck BMD and lumbar spine BMD. In the present study, we observed low frequency of C-allele in LLIs, indicating lower risk of skeletal and bone related problems. The other allele (T-allele), which is also found to be associated with decreased body height [43], is in high frequency in LLIs. Multiple research studies found a negative correlation between height and longevity. Shorter people tend to have higher resistance to chronic diseases, especially in middle age [44].

Anemia is also one of the major health concerns in older adults. It is associated with cognitive deficits and reduced physical performance. Several genes and genetic variants are known to affect iron levels, hemoglobin levels and anemia. ACO1 is one such gene that encodes a bifunctional protein which controls the levels of iron within the cells. We observed that the variant in this gene, rs10970985, has lower frequency of C allele in LLIs compared to controls. Further research studies are required to understand the role of this variant.

HSPA5 gene (heat shock protein family A (Hsp70) member 5) encodes immunoglobulin protein that is an essential component of folding/unfolding processes and regulation of Ca2+ homeostasis in the endoplasmic reticulum (ER) [45]. Disturbed homeostasis leads to ER stress, causing inflammation, metabolic disorders and neurodegenrative diseases. It is also associated with anxiety disorders such as depression and post-traumatic stress disorder (PTSD) [46]. Research studies found T-allele of rs391957 from HSPA5 gene to be a protective factor for anxiety and neuroticism [29-30]. In the present investigation we observed the frequency of T-allele to be higher in LLIs compared to younger controls, indicating that lesser anxiety leads to increased lifespan.

ABCC2 (ATP binding cassette subfamily C member 2) gene encodes a protein called multidrug resistance protein 2 (MRP2) which is involved in metabolism and clearance of certain drugs from organs and tissues. It is also known to play a major role in biliary transport. Mutations in ABCC2 gene were observed in patients with Dubin-Johnson syndrome, an autosomal recessive disorder characterized by conjugated hyperbilirubinemia. T allele of rs2002042 from ABCC2 gene was found to be associated with intrahepatic cholestasis of pregnancy and nonalcoholic fatty liver disease [47,31]. In our study, we observed lower frequency of T allele in LLIs compared to controls, indicating undisturbed biliary transport, good enterohepatic circulation of bile acids and healthy gut leading to longer lifespan.

Carriers of G allele at rs365990 have 1.725 times higher chances of surviving to older age. Previous studies indicate that G allele of rs365990 is associated with decrease in heart rate [32-33]. Low resting heart rate results in increased lifespan due to reduced stress on arterial cells, reduced stiffening of aorta and reduced cardiovascular abnormalities [48-49]. Faster heart rate is related to high metabolic rate, development of free radicals, oxidative stress and faster aging. Studies also indicate that heart rate lowering through medication or selective sinus node inhibition increases the lifespan.

Our pathway enrichment analysis confirmed that the molecular processes associated or pathways with longevity are significantly different in LLIs. Pathways with the genes (through SNPs) involved in oxidative stress, apoptosis, DNA damage repair, glucose metabolism and energy metabolism are differentially regulated in LLIs. IRS2 and PRKAA1 genes that are related to regulation of insulin secretion and glucose levels are common across most of the significant pathways. Several previous studies indicated the role of PI3K-Akt signaling pathway in ageing [50, 51]. In addition to that, our study revealed pathways related to non-alcoholic fatty liver disease (NFLD), inflammatory bowel disease (IBD) and inflammation to be associated with ageing. Screening the variants associated with these disorders early on in life helps in recommending preventive measures, and in promoting healthy ageing.

Major limitation in our study is the sample size. Increasing the sample size will increase the statistical power of the study, and may identify additional variants that increase our understanding of factors affecting longevity. Also, gender-specific analysis is not feasible due to small sample size. But our results can provide some indication on variants and factors associated with longevity in the population, and help in further follow-up studies with larger sample sizes. Additional information on clinical and lifestyle variables, such as BMI, cholesterol levels, diet patterns, physical activity, smoking and alcohol consumption help in identifying the variants associated with interaction of those variables with longevity.

Allele frequencies of the longevity-associated SNPs vary among populations studied under 1000 Genomes project (Supplementary Table 3). Major alleles in one population were observed to be minor alleles in other populations. For example, G allele of rs3903239 associated with atrial fibrillation is a minor allele in SAS, but major allele in EAS (East Asians). Similarly, alleles associated with longevity from the SNPs of FOXO3 gene have in general higher frequency in AFR and SAS populations. This indicates the need for replication studies in different populations to confirm the associations in each population.

In conclusion, our study identified genetic variants associated with longevity in Indian population that is underrepresented in genetic research. Clinical and lifestyle factors inferred through the identified variants can guide in recommending strategies for healthy aging and longevity at population-wide and an individual level.

## Data Availability

Data is available to researchers on collaboration with Mapmygenome India Limited by contacting

## Notes

### Competing Interest Statement

The authors have declared no competing interest.

### Funding Statement

This study did not receive any funding

### Author Declarations

Data was taken from GenomegaDB, a genetic database available at Mapmygenome

